# Monitoring social distancing and SARS-CoV-2 transmission in Brazil using cell phone mobility data

**DOI:** 10.1101/2020.04.30.20082172

**Authors:** Silvano Barbosa de Oliveira, Victor Bertollo Gomes Pôrto, Fabiana Ganem, Fabio Macedo Mendes, Maria Almiron, Wanderson Kleber de Oliveira, Francieli Fontana Sutile Tardetti Fantinato, Walquiria Aparecida Ferreira de Almeida, Abel Pereira de Macedo Borges, Hector Natan Batista Pinheiro, Raíza dos Santos Oliveira, Jason R. Andrews, Nuno R Faria, Marcelo Barreto Lopes, Wildo Navegantes de Araújo, Fredi A. Diaz-Quijano, Helder I. Nakaya, Julio Croda

**Author notes:** These authors contributed equally to this work (Silvano Barbosa de Oliveira, Victor Bertollo Gomes Pôrto).

## Abstract

Social distancing measures have emerged as the predominant intervention for containing the spread of COVID-19, but evaluating adherence and effectiveness remains a challenge. We assessed the relationship between aggregated mobility data collected from mobile phone users and the time-dependent reproduction number R(t), using severe acute respiratory illness (SARI) cases reported by São Paulo and Rio de Janeiro. We found that the proportion of individuals staying home all day (isolation index) had a strong inverse correlation with R(t) (rho<-0.7) and was predictive of COVID-19 transmissibility (p<0.0001). Furthermore, indexs of 46.7% had the highest accuracy (93.9%) to predict R(t) below one. This metric can be monitored in real time to assess adherence to social distancing measures and predict their effectiveness for controlling SARS-CoV-2 transmission.

**One Sentence Summary:** Mobility data to monitoring social distancing in the COVID-19 outbreak

## Main Text

The coronavirus diseases 2019 (COVID-19) pandemic has caused more than 2,900,000 cases and 200,000 deaths worldwide as of April 26, 2020(*1*). In the absence of vaccines and effective pharmacological interventions, social distancing measures are critical to mitigate the impact on healthcare systems and allow time for a public health response around the globe(*2, 3*).

There is growing evidence that a significant reduction in new locally-transmitted cases have been achieved in Asia and Europe following restrictions on urban mobility and travel(*2–6*). To reduce the peak of transmission, starting in March of 2020, the Brazilian states of São Paulo and Rio de Janeiro in Brazil, with a population of 63 million people, implemented non-pharmacological interventions and recommended social distancing measures. This implied reductions in public gatherings; the closure of schools, universities, and businesses such as restaurants, bars and gyms; and the institution of remote or virtual work for older adults and individuals with underlying medical conditions. In light of these public health interventions only essential services (i.e., grocery stores, emergency health service) remained open. To prevent the expansion of COVID-19 epidemic, the non-pharmacological measures have to bring the time-dependent reproduction number (R(t)) to less than 1. This requires a 50–60% reduction of a baseline R(t) between 2 and 2.5(*2*).

Using data from hospitalization due to severe acute respiratory illness (SARI) as a proxy for severe cases of COVID-19, we recently reported an important decline in the R(t) in the metropolitan area of São Paulo after the implementation of social distancing measures(*7*). Although R(t) is a useful tool to monitor the epidemic, it is subject to significant delays in notification. In Brazil, a time lag of 8 days exists between the date of onset of symptoms and the date of notification of a SARI case(*8*). Given this time, relying solely on the measurement of R(t) bears a likely risk of introducing significant delays in detecting changes in the epidemic dynamics. In an epidemic with cases doubling in 5.2 days(*9*), a delay of 1–2 weeks in the implementation of effective social distancing would overwhelm the capacity of the healthcare system, resulting in a growing number of excess deaths(*10–13*). Thus, a timelier metric for COVID-19 transmission potential is required to evaluate social distancing interventions.

The wide-ranging usage of mobile phone provides an unique opportunity to monitor and control the spread of infectious diseases(*14–17*). Mobile phones with geolocation enablement can track movement in a timely and precise manner. To our knowledge, no study thus far has assessed the use of mobile phones for this purpose in Brazil. Monitoring human activity through mobile aggregated data can permit measurement of the effectiveness of social distancing measures in reducing transmissibility of the SARS-CoV2 virus(*18*). Nonetheless, the association between urban mobility and COVID-19 transmissibility has not yet been established. In this study, we assessed the relationship between social isolation measures, by using human urban movement data and the transmissibility of COVID-19.

From February 1st to April 10th, 2020 a total of 21,426 hospitalizations in 853 hospitals (58%) due to SARI were reported in the São Paulo (SP) state. A clinical diagnosis of COVID-19 was confirmed by molecular testing for 4,111 (19% of total SARI cases), 16,892 (79%) remained as suspected COVID-19 cases and 423 (2%) were confirmed for other respiratory virus diseases in SP state. During the same period the RJ state had reported 2,540 hospitalizations in 320 hospitals (62%) due to SARI, 402 (16%) were confirmed by molecular testing for COVID-19, 2,090 (82%) remained as suspected and 48 (2%) have been confirmed for other respiratory viruses (Fig. 1A and 1D). Most SARI cases in SP and RJ have not been tested due to the shortage of SARS-CoV2 real-time polymerase cycle reaction (RT-PCR) diagnostic screening. In 2019, during the same period, 1,550 and 220 SARI cases were reported in SP and RJ, respectively. This corresponds to 1,282% and 1,054% increase in SARI cases in SP and RJ respectively between the same periods in 2019 and 2020.

### Implemented social distancing Measures

Starting on March 13, 2020, two days after the World Health Organization declared COVID-19 as a pandemic, the states of São Paulo and Rio de Janeiro implemented a series of non-pharmacological interventions. These measures were implemented gradually in both states, some of these interventions are described on **Figure 1**. To investigate the impact of social distancing measures in SARS-CoV-2 transmission, we compared daily aggregated mobility data for São Paulo and Rio de Janeiro with *R* estimates obtained from SARI data. Here, we calculate an “isolation index” as a ratio between the number of people staying at home on a given day divided by the number of cell-phone users living in the state or city (see **Material and Methods**). First, we find that the mean isolation index from February 1st to April 10th was 40.2%, ranging from 18.5% to 69.4%. In São Paulo state, mean isolation index ranged from 13.5% to 67.9% and in the Rio de Janeiro State from 16.6% to 69.4%, with no significant difference observed between the two states (p-value = 0.210). Second, after the start of interventions we observe a sharp and significant increase in the isolation index and a corresponding decrease in the R(t) (**Figs. 1B** and **1E**). Overall, we find that temporal periods with R(t) ≥1 had a mean isolation index below 29.3% (SD = 8.2%) while those with R(t) <1 had a mean isolation index above 53.4% (SD = 3.0%) (**Figs. 1C** and **1F**).

**Fig. 1.**
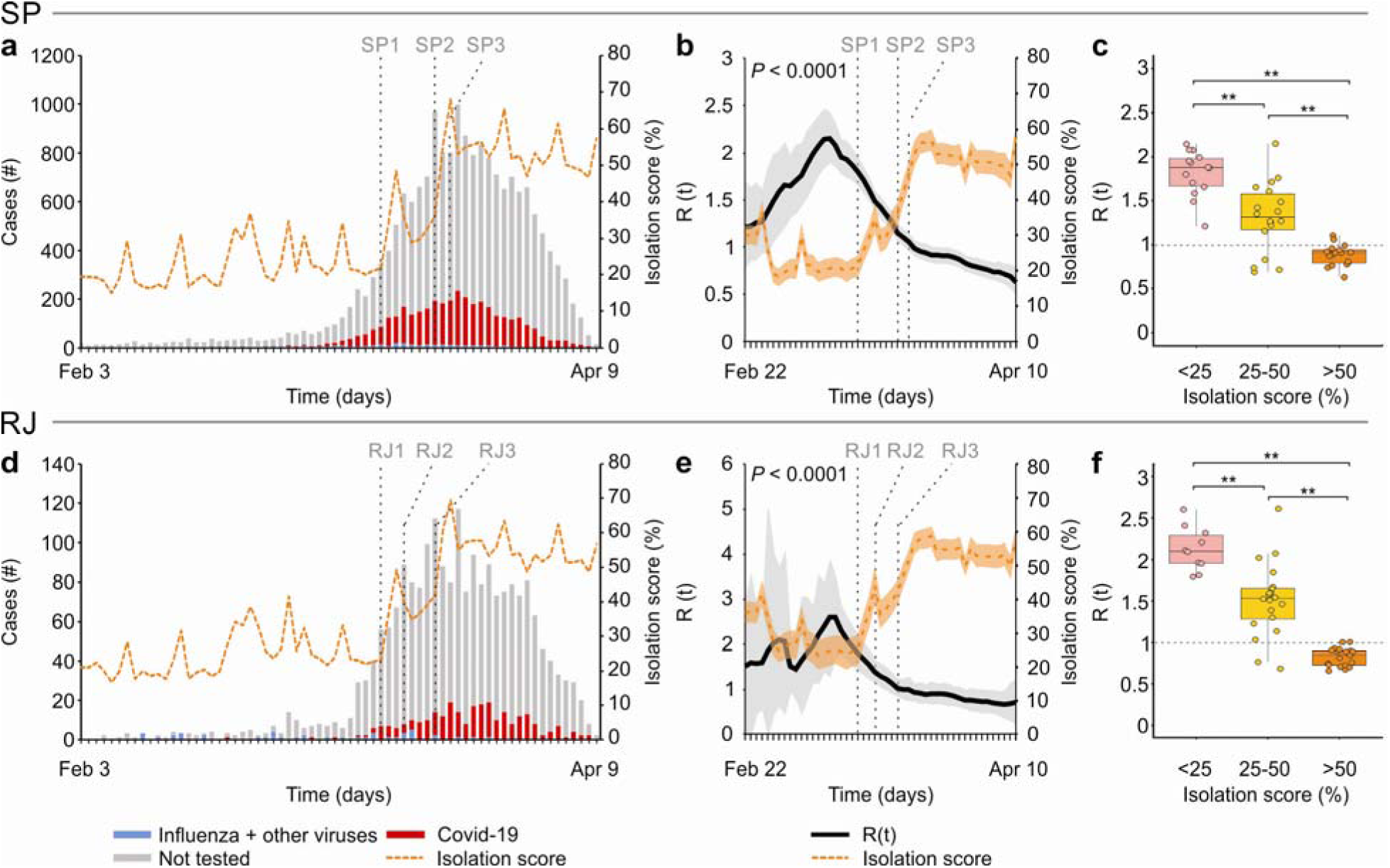
Impact of interventions in the reduction of severe acute respiratory transmission São Paulo and Rio de Janeiro states, Brazil. A and D: epidemic curve of SARI cases by date of onset of symptoms for São Paulo and Rio de Janeiro, respectively; B and E: Daily Time dependent reproductive number (R(t)) (grey) and isolation index (orange) (curves smoothed by Kalman Filtering method), SP and RJ respectively; C and F: boxplot showing the median, interquartile range and range of the association between of R(t) and isolation index categories. SP1: March 13, 2020, SP recommended reduction in public gatherings events and closure of schools; SP2: March 20, 2020, SP declares Public Calamity and prohibits religious ceremonies; SP3: March 22, SP establishes quarantine in the entire state, closure of night clubs, shopping centers, gyms, bars, restaurants and bakeries; RJ1: March 13, 2020, RJ recommended remote work, suspended public gathering events, closed schools, and entertainment establishments, prohibited the access of visitors to prisons and to COVID-19 hospitalized patients; RJ2: March 16, 2020, RJ prohibited the access to tourist places, including beaches and public pools, restricted public transport such as bus lines, airlines and cruise ships coming from states or countries with COVID-19 circulation, closed bars and restaurants, closed gyms, shopping centers and similar establishments and set restrictions on public transportation; RJ3: March 20, 2020, RJ declared Public Calamity due to the COVID-19 pandemic

### Mobility data predicted the time-dependent reproduction number R(t)

We next conducted cross-correlation analyses with different lag days and the Granger test to investigate if the isolation index measure obtained from mobility data is able to predict R(t). After the intervention began, it was also observed a gradual decrease in R(t) (Fig. 1B and 1E). Furthermore, using SARI cases, cross-correlation analyses showed that isolation was highly correlated with R(t) (rho<-0.7) in a lag period of up to five days in SP and RJ. Considering only COVID-19 confirmed cases, the R(t) and isolation index were moderately correlated (rho >-0.7) using a lag period from -2 to 0 (Fig. 2A and B).

**Fig. 2.**
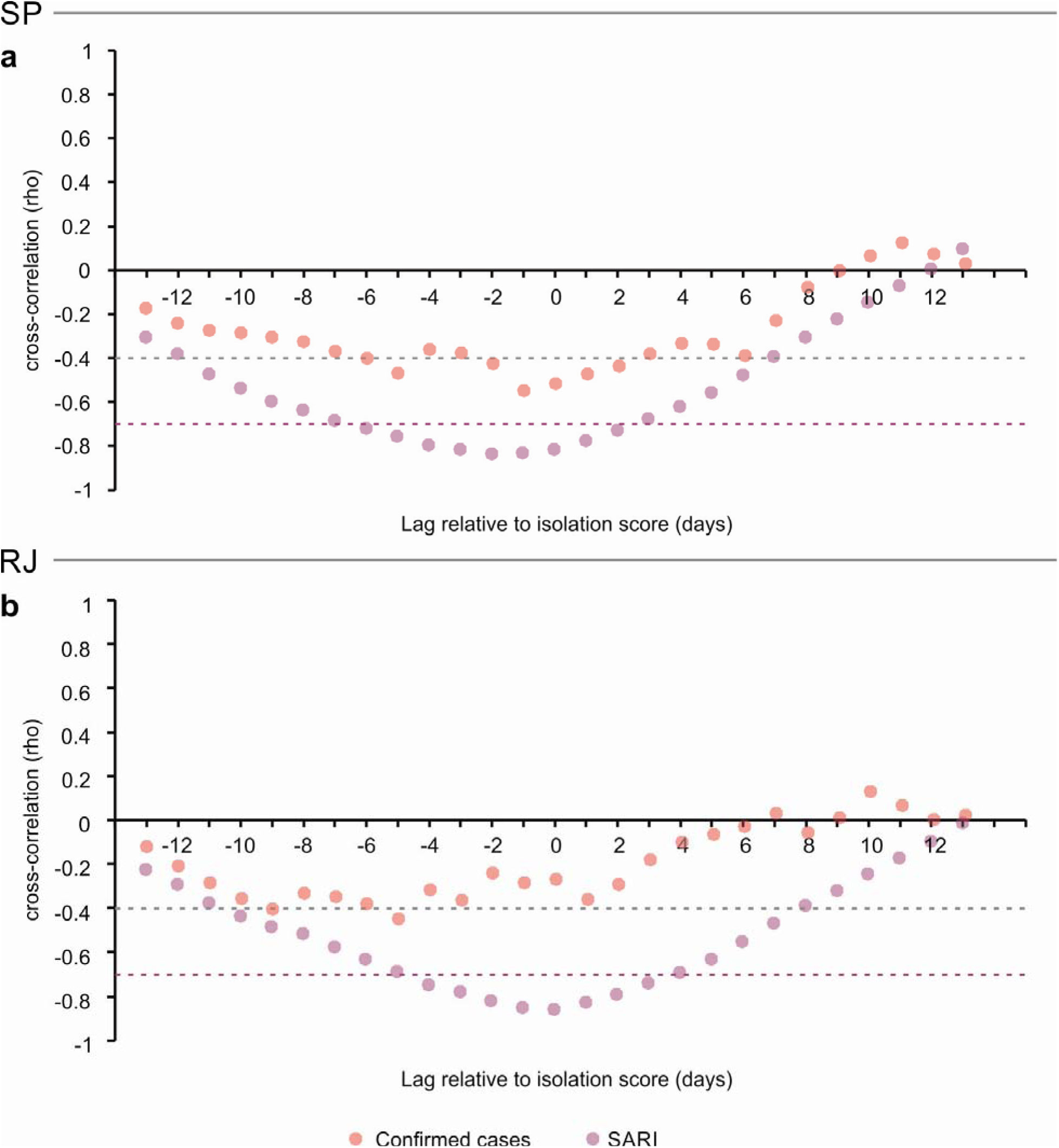
Cross-correlation (rho) between isolation index and reproductive number (R(t) estimated using SARI (purple circles) and confirmed COVID-19 (red circles) cases in São Paulo (A) and Rio de Janeiro (B).

The isolation score exhibited an area under the ROC curve 93.8% (95% CI: 88% - 99.5%) to predict *R* values under 1.0 (**Fig. 3A**). We obtained the best accuracy with score values close to 50%. Particularly, we observed the highest accuracy (93.9%) with the cut-off of 46.7%, and cutoffs higher than 50% exhibited specificity greater than 93% (**Fig. 3B**). Furthermore, 88.9% of the areas with at least 50% isolation score had an *R* <1. In contrast, 90.3% of the observations with an isolation score <50% had an *R* ≥1. The isolation scores, for both states combined, when greater than 50%, were associated with a mean *R* of 0.9 (ranging from 0.6 to 1.1) (**Fig. 1C and 1F**). Scores between 25–50% had a mean *R* of 1.4 *R* (ranging from 0.7 to 2.6). On the other hand, isolation scores lower than 25% exhibited a mean *R* of 1.9 (ranging from 1.2 to 2.6).

**Fig. 3.**
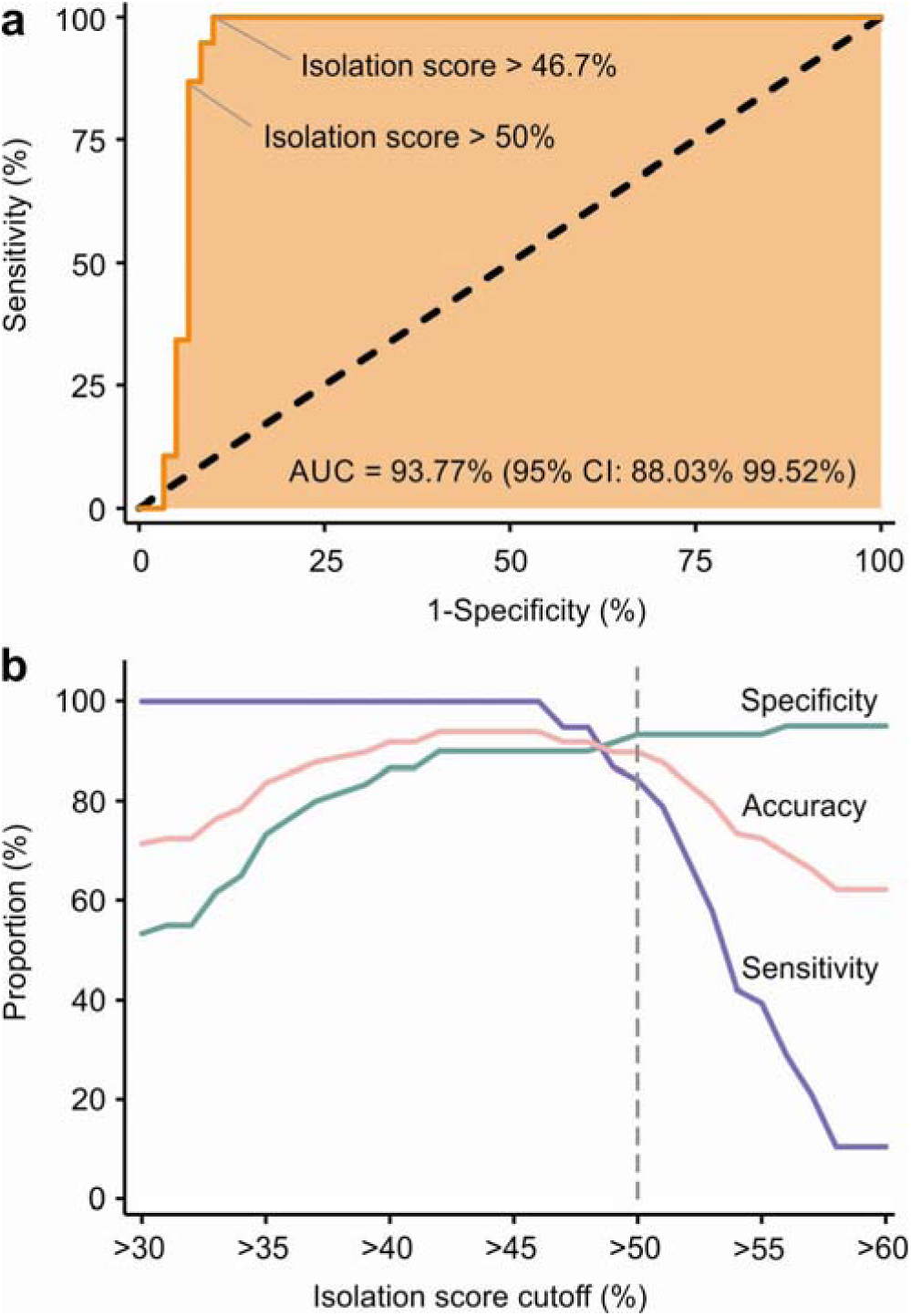
A. Receiver operating characteristic curve of isolation index for prediction of R(t)<1. B. Sensitivity, Specificity and Accuracy of isolation index cut-offs to predict R(t)<1.

To investigate the effectiveness of interventions in areas with different human development indexes (HDI), we analyzed the isolation score time series for the 10 largest cities in the states of São Paulo and Rio de Janeiro stratified by HDI(*19*). We found no significant difference between the groups (**Fig. 4**). To validate the findings, we aggregated 20 cities in two groups stratified by HDI (lower and higher values). We observed the same trend and strong correlation, regardless of the group (higher and lower HDI) (**Fig. S1A**). Interestingly, we found an identical cut-off point of isolation index (50%) which was correlated with R(t) values below 1 (**Fig. S1B**).

**Fig. 4.**
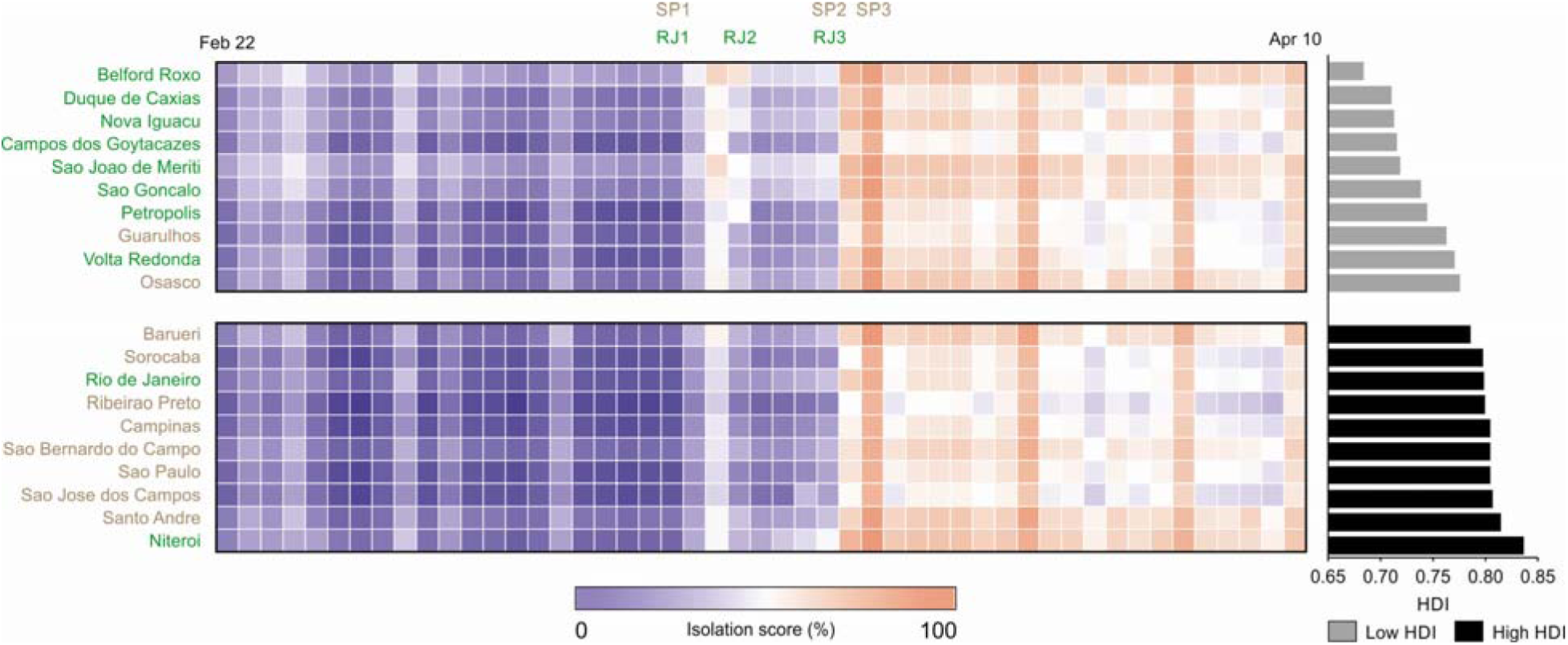
Isolation index time series for the 10 largest cities in the states of São Paulo (SP) and Rio de Janeiro (RJ). SP1, SP2, SP3, RJ1, RJ2 and RJ3 correspond to intervention dates (See Fig. 1 legend)

### Simulation of different interventions scenarios with isolation indexs above 50%

Assuming the R(t) values associated with isolation indexs above 50% we next simulated different lengths of social distancing interventions, 30, 60 and 90 days (see **Materials and Methods**). An intervention of only 30 days was able to reverted the intensive care units (ICU) demand curve only in those scenarios with the lowest values of R(t), and was also associated with a quick rebound of the ICU demand curve far above the existing capacity (Figure 5). Prolonging the intervention to 60 days managed to revert the curve in most scenarios and delayed the second epidemic wave. An intervention of 90 days substantially delayed the second epidemic wave in all scenarios, although it had minimal impact on the height of both curves. According to our simulation, Rio de Janeiro state is able to stay below the ICU bed capacity threshold in the first epidemic wave in all scenarios. Conversely, São Paulo state needs to steadily increase its capacity in order to manage the first epidemic curve. Lower values of R(t) were associated with smaller epidemic waves during the intervention but higher second epidemic waves (Figure 5).

**Fig. 5.**
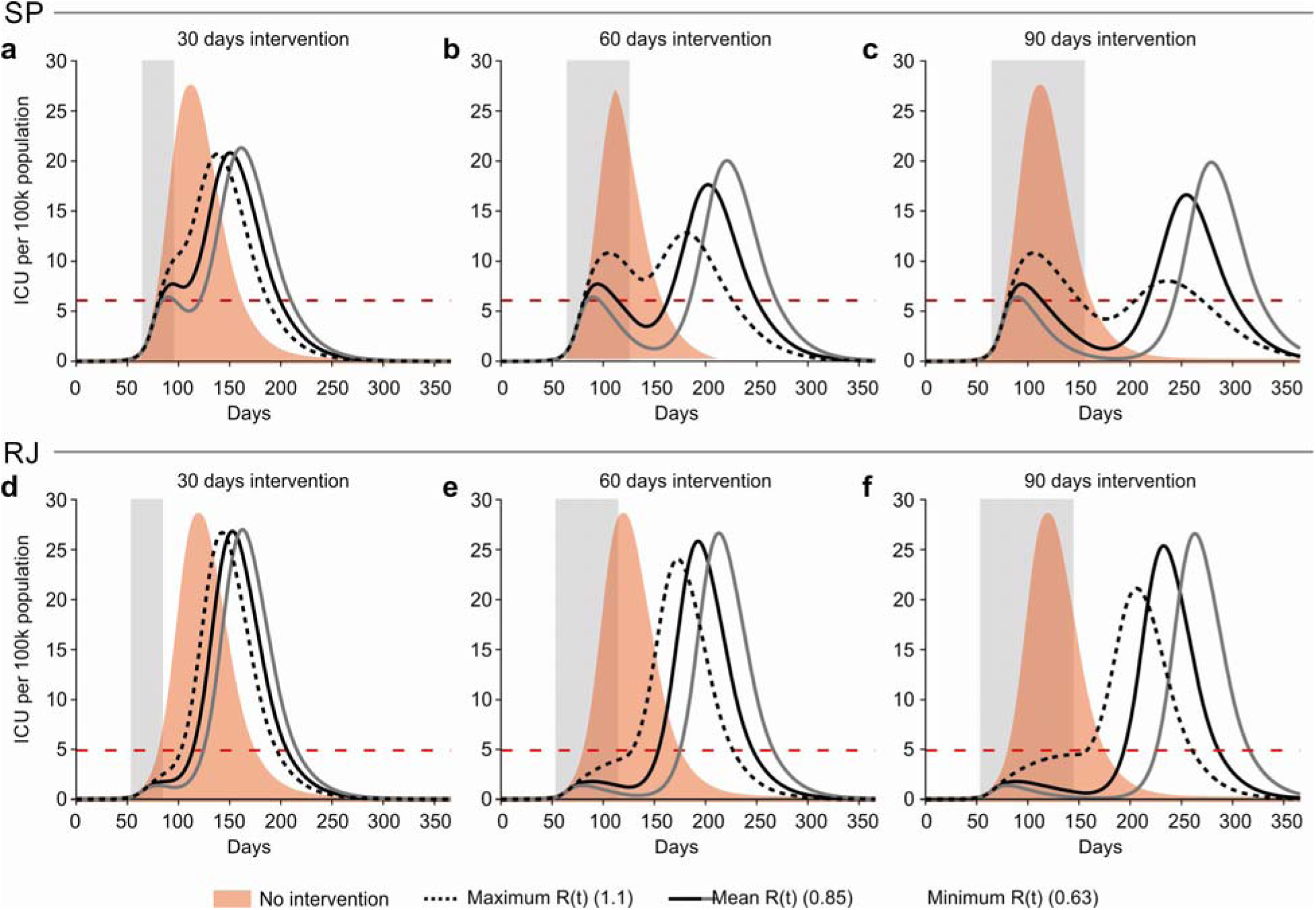
Simulations of intensive care unit (ICU) demand according to different lengths of social distancing interventions for the states of São Paulo (top row) and Rio de Janeiro (bottom row) A, B and C: simulations for the São Paulo state assuming *R* values associated with isolation scores above 50% during 30, 60 and 90 days of intervention respectively; D, E and F: simulations for the Rio de Janeiro state assuming R(t) values associated with isolation scores above 50% during t 30, 60 and 90 days of intervention respectively. For each scenario we plotted the mean, minimum and maximum R(t) values in the >50% isolation score category. The red horizontal line corresponds to ICU capacity for each state.

## Discussion

Individual-based control efforts of COVID-19 can be challenging to implement due to the high infectiousness, relatively mild and moderate symptoms and pre symptomatic transmission^23–25^. Consequently, social distancing measures are required to control COVID-19 in most scenarios. Here, we demonstrated that the initial social distancing measures adopted by the states of São Paulo and Rio de Janeiro successfully reduced R(t) < 1 by decreasing mobility across populations with lower and higher development index. We further demonstrated a strong correlation between the social isolation index and R(t), and showed that isolation indexs above 50% lead to R(t)<1 in most cases (89%). Our findings suggest that the isolation index can be used to monitor the effectiveness of social distancing measures and guide further interventions. By using aggregated mobility data, public health officials and policymakers can monitor in real time regional differences in social distancing intervention effectiveness and propose specific actions to reduce the transmission in specific locations(*20*).

Individual-based control efforts for COVID-19 can be challenging to implement due to the high infectiousness, relatively mild and moderate symptoms and pre-symptomatic transmission (*21–23*). Consequently, social distancing measures are required to control COVID-19 in most scenarios.

The structural impacts of social distancing should also be considered while managing its impact on disease transmission and, consequently, the healthcare systems’ capacity. A previous analysis demonstrated that social distancing adopted during the Spanish flu pandemic was associated with better economic outcomes(*24*). However, society dynamics have changed dramatically during the past 102 years, and a continued monitorization of the COVID-19 socioeconomic impact is urgently needed. Suboptimal interventions for COVID-19 could be potentially catastrophic to public health and lead to a significant number of deaths. Thus, finding the optimal “therapeutic dosage” of such interventions is crucial, and requires a reliable and timely tool to monitor its effects.

R(t) depends on the contact-rate among individuals; but the frequency of these contacts is difficult to quantify in real time through conventional approaches(*25*). We observed an inverse relationship between R(t) and the isolation index for São Paulo and Rio de Janeiro States, the two mostly affected by the epidemic. We find that increases in the isolation index from lower than 25% to greater than 50% have lead to a reduction of R(t) from approximately 2 to values less than 1, such as we observed. Highly-effective social distancing could reduce COVID-19 transmission enough to make a strategy based on contact tracing feasible, as is taking place in South Korea and Singapore(*26, 27*).

By using the R(t) values associated with isolation indexs above 50%, we simulated social distancing interventions with varying time extensions. In most scenarios, interventions lasting between 30 to 60 days would blunt the first epidemic wave, although a shorter intervention led to a quick rebound and was not sufficient in scenarios with higher values of R(t) during the intervention. Lower values of R(t) resulted in a more effective control during the first wave, but with a caveat that the second epidemic wave was became significantly larger (albeit smaller than the initial peak that would have been otherwise seen in the absence of interventions). Furthermore, the delay in this peak allows the preparation of healthcare systems to mitigate health impacts by securing equipment and supplies, bolstering ICU capacity, planning for personnel needs and implementing infection control policies.

Higher values of R(t) during the intervention led to more evenly distributed epidemic waves, which would make it possible to implement less stringent interventions during following waves. Although it is tempting to propose that social distancing interventions that would lead to an R(t) value of 1.1, such a strategy could have potentially catastrophic consequences. The reason for this is that values of social indexs below 50% are associated with a mean R(t) value of 1.4 (ranging from 0.7 to 2.6), which was not significantly different from no intervention. Aiming at achieving a R(t) value of 1.1 by lowering the social isolation index could therefore lead to a scenario similar to the natural history of COVID-19. It is thus more effective and probably safer to set a goal of R(t) below 1 and implement social distancing interventions that lead to social isolation indexs above 50%. This will require close monitoring after the intervention is relaxed, since it is very likely that a second intervention will be needed to flatten a second epidemic wave.

The findings of our simulations need to be interpreted cautiously. We did not aim to precisely replicate the epidemic in both states but merely to simulate hypothetical scenarios, exploring different strategies. Also, our simulations are heavily influenced by the assumptions and parameters that were built into the model. A key assumption in our model is that infection leads to persistent immunity after recovery. Data on SARS-CoV-2 immunity is scarce, preliminary data on rhesus macaques have demonstrated immunity after an initial infection(*28*, *29*) and plasma from recovered individuals was tested to neutralize the virus *in vitro* and *in vivo*(*30–33*). On the other hand antigenic drifts(34) as well as waning immunity might lead to the loss of herd immunity. If immunity is not long lasting, as has been estimated for other coronaviruses such as HCoV-OC43 and HCoV-HKU1, COVID-19 will likely enter a regular transmission cycle and become endemic(*35*).

Another aspect that needs to be taken in consideration is that we simulated epidemics in entire states, assuming homogeneous transmission within the state. However, it is possible that different cities will have non synchronized COVID-19 epidemics, hence interventions will need to be tailored for each city or metropolitan area individually. Moreover, our assumption that only 50% of the established ICU capacity could be used for COVID-19 might be an underestimation of the total capacity, given that during the epidemic efforts have been made to increase hospital capacity in both states, which could lead to the need of less stringent interventions.

Another potential source of bias is that we assumed that the reporting rates were stable during the epidemic. If reporting rates of SARI cases increased during the initial stages of the epidemic our initial R(t) estimates would have been overestimated. Further, if the reporting rates are decreasing, it could mean that the decrease in R(t) that was observed would be partially a result of a reporting bias. Although Brazil has had changes in the reporting systems for COVID-19 during the epidemic, the dataset which was used is based solely on hospitalized SARI cases which required hospitalizations. However, we used SIVEP-GRIPE data, a stable and a well-established system and recommendations for its use have not changed during the epidemic, we believe that this is the most reliable and consistent source of data for severe cases of COVID-19 in Brazil. We also accounted for potential bias related to reporting delays, which affect the end of a time series, by applying a correction factor which corresponds to the inverse of the probability of being reported up to the last date of data being collected.

Another limitation of our approach is that mobility data from mobile phones are a coarse measure of physical distancing, which does not directly capture changes in the number, duration or character of human interactions. Additionally, this approach does not account for other behavioral changes in the population (such as hand washing, respiratory etiquette and universal mask usage) that could also alter transmission patterns and lead to changes in R(t).

Our findings underscore the importance of early implementation of social distancing measures to reduce SARS-CoV-2 transmission. The strong association observed here indicates that urban mobile phone-derived isolation scores are able to track temporal fluctuations in *R* during social distancing interventions in Brazil. If additional behavioral changes are undertaken and sustained, the isolation score could provide a real-time metric to assess effectiveness of interventions. A major contribution of our approach is that the social isolation index data is readily available on a daily basis, in contrast with the R(t) measurement, which is subject to delays (up to two weeks). Using this index will allow for a more timely assessment of the epidemic dynamics and for planning of public health mitigation strategies.

## Data Availability

Mobility data is available free of charge for non-private organizations after contacting the Inloco company and signing a term of responsibility. The SIVEP-GRIPE csv dataset is available at https://opendata.saude.gov.br/dataset/notificacoes-srag-2019. All other data that support the figures within this paper and other findings of this study are available at https://github.com/pydemic/covid-19.

## Acknowledgments

We thanks the Department of Immunizations and transmissible diseases to support this work and In Loco company to provide the isolation index database available for free. Acknowledgments follow the references and notes but are not numbered. **Funding:** FADQ and JC were granted a fellowship for research productivity from the Brazilian National Council for Scientific and Technological Development – CNPq, process/contract identification: 312656/2019–0 and 310551/2018–8, respectively. NRF is funded by a Medical Research Council and CADDE partnership award (MR/S0195/1) and by a Sir Henry Dale Fellowship from Wellcome Trust and Royal Society (204311/Z/16/Z). **Author contributions:** M.B.L., W.N.A., F.A.D., H.I.N and J.C. designed the research; B.O, V.B.G.P., F.G., F.M.M., F.A.D. and H.I.N performed the research; B.O, V.B.G.P., F.G., F.M.M., M.M, W.N.A., F.A.D., H.I.N and J.C. analyzed the data; and S.B.O, V.B.G.P., F.G., F.M.M., M.M, W.K.O., F.F.S.T.F., W.A.F.A, J.R.A., N.R.F., M.B.L., W.N.A., F.A.D., H.I.N, J.C. wrote and edited the paper. **Competing interests:** Authors declare no competing interests. **Data and materials availability:** Mobility data is available free of charge for non-private organizations after contacting the Inloco company and signing a term of responsibility. The SIVEP-GRIPE csv dataset is available at https://opendata.saude.gov.br/dataset/notificacoes-srag-2019. All other data that support the figures within this paper and other findings of this study are available at https://github.com/pydemic/covid-19.

## Ethical approval

This study followed Brazilian and International legislation for conducting human research. This research project was approved by the National Research Ethics Committee (Comissão Nacional de Ética em Pesquisa, CONEP) in Brazil, Register number (CAAE): 11946619.5.0000.5421.

## Material and Methods

### Mobility data

We used geolocation data collected between February 6, 2020 to April 10, 2020. Urban mobility patterns were extracted from unidentifiable personal information. We settled the likely home-residence location based on the place most often visited during non-working hours. Device location data was compiled and aggregated at both city and states level by the technology company *In Loco* (São Paulo, Brazil). As of February of 2020, Brazil totaled more than 227 million mobile phones devices in use with the states of São Paulo (66,939,000) and Rio de Janeiro (18,633,000) ranking first and third with the most devices in the country. In the same month, the technology company *In Loco* (São Paulo, Brazil) accessed geocoded information from 17,482,780 million users in the state of São Paulo (26%) and 5,390,130 (29%) in the state of Rio de Janeiro, and it reported more than 700 million monthly geolocations tracked to physical locations in these two states.

An isolation index was calculated as a ratio between the number of people staying at home on a given day divided by the number of mobile phone users living at the same state or city, expressed as a percentage. Only those mobile phones with geolocation app tracking active were considered for the index calculation.

### Severe Acute Respiratory Illness and time-dependent reproduction number

Brazil has a well-established surveillance system for severe acute respiratory illness (SARI), and notification of SARI cases has become mandatory since 2009. Since then hospitalized SARI cases as well as cases of influenza like illness (ILI) reported by the sentinel surveillance sites are registered in an electronic database, SIVEP-GRIPE. Although Brazil initially established a reporting system for mild COVID-19 cases based on the REDCap platform, this was shifted on March 25, 2020 to a new reporting system, e-SUS VE. At the same time, input of SARI cases into SIVEP-GRIPE database has remained consistent. We used all hospitalized SARI cases which were reported by the states of São Paulo (SP) and Rio de Janeiro (RJ) from the same period of geolocation data collected. We included SARI cases which were confirmed for COVID-19 (positive RT-PCR for SARS-CoV-2 in any respiratory sample), as well as those suspected without any etiological diagnosis. We excluded cases with a positive PCR for other respiratory viruses and that also had not been tested for COVID-19 or that tested negative. Due to the lack of massive testing, the inclusion of SARI cases without an etiological diagnosis was carried out assuming that most SARI cases were expected to be related to COVID-19 during the epidemic. The exclusion of these cases would have led to biased results due to delays in ascertainment of COVID-19 cases. The epidemic curve was constructed using the date of onset of symptoms.

To correct for reporting delays, we applied a daily correction factor based on the observed delays between the date of onset of symptoms and the date of data reporting. This weight corresponded to the inverse of the probability of being reported up to the last date of data being collected(*18*). Moreover, we restricted the analysis to patients who developed symptoms until 4 days before the last data entry. The R(t) was calculated considering the epidemic curve and serial interval observed in the COVID-19 cases in Brazil, using the R0 package available in *R*(*19*). To test the temporal relationship between the isolation index and R(t) time series, we used cross correlation analysis with various lag days and the Granger test (isolation index predicting R(t)).

We summarized the ability of the isolation index to predict a R(t) lower than 1, by calculating the area under the ROC curve (AUC). Moreover, we looked for the cut-off point with the highest accuracy (proportion of observations with R(t)<1 and R(t)≥1, respectively above and below the index cut-off point) and supported a recommendation by rounding up that isolation index point.

We additionally compared the R(t) for the isolation index categories (under 25%, 25 - 50% and greater than 50%, based on ROC curve) using Kruskal Wallis test and post-hoc analysis was conducted through the Dunn test for multiple comparison. We also stratified the analysis by Human Development Index (HDI) in order to assess for an interaction with socioeconomic status. For this analysis, we used data from the 10 largest cities in the states of São Paulo (SP) and Rio de Janeiro (RJ) and evaluated changes in the trends of their indexs, split into two groups: high HDI (range from 0.786 to 0.837) and low HDI (range from 0.684 to 0.776) and compared the isolation indexs from February 22, 2020 to April 10, 2020 between the groups(*20*).

### Application in intervention-effect modeling

To simulate different interventions scenarios we used an age stratified SEIR model which includes compartments for individuals requiring hospitalization and intensive care (*6*). We considered three different time frames of social distancing interventions (30, 60 and 90 days) and simulated the prevalence rate, ICU demand and number of deaths for COVID-19. During the intervention we used the mean, maximum and minimum reproductive numbers (R) associated with isolation indexs greater than 50% which were observed in our dataset. We used a Basic Reproductive Number (R0) equal to 2.27 at the beginning of the epidemic, which was the peak value for the observed R(t) series. After intervention we used an R value of 1.8, assuming that after ceasing the interventions residual changes in the population’s behavior (such as mask usage for example) would lead to approximately a 20% reduction in transmissibility. The remaining model parameters are described in Table 1.

In all simulations, the model was seeded with a prevalence of 10^−7^ per population, which corresponds to 4.4 cases in SP and 1.65 cases in RJ, and evolved with R0 = 2.27 until reaching the cumulative mortality rate which was observed at the beginning of the intervention in each state. After reaching the established threshold, the intervention starts in our model, at that point the reproductive number gradually changes during 8 days until reaching the pre specified R value associated with the intervention. This 8 days transition period was chosen based on the observed delay between the first intervention and reaching isolation indexs above 50% in both states. With this assumption, we wanted to compare how both states would evolve and how changes in demography, capacity of the healthcare systems and the timing of the intervention would influence the ability to cope with the epidemic under otherwise similar conditions. Hospital and ICU capacities were collected from the National Registry of Health Establishment (CNES). In SP and RJ, 1481 and 523 hospitals were active in the month of February of 2020, respectively. The model assumes that 50% of the total number of ICU beds available in the public healthcare system could be allocated to COVID-19.

**Table S1.** Parameters used in the age stratified SEIR model to forecast the ICU beds demand, prevalence and deaths.

| Parameter | Value | Source |
| --- | --- | --- |
| R0 | 2.270 | Ganem et al(6) |
| R(t) during intervention - min | 0.628 |  |
| R(t) during intervention - mean | 0.852 |  |
| R(t) during intervention - max | 1.106 |  |
| R(t) after intervention | 1.800 |  |
| Incubation period | 3.69 days | Li et al(36) |
| Infectious period | 3.47 days | Li et al(36) |
| Proportion of infected individuals<br>hospitalized / Infection fatality ratio (IFR)<br>by age group |  | Verity et al(37) |
| 0 - 9 years | 0.00% / 0.00161% |  |
| 10 - 19 years | 0.0408% / 0.00695% |  |
| 20 - 29 years | 1.04% / 0.0309% |  |
| 30 - 39 years | 3.43% / 0.0844% |  |
| 40 - 49 years | 4.25% / 0.161% |  |
| 50 - 59 years | 8.16% / 0.595% |  |
| 60 - 69 years | 11.8% / 1.93% |  |
| 70 - 79 years | 16.6% / 4.28% |  |
| >80 years | 18.4% / 7.80% |  |
| Case fatality in individuals requiring intensive care (CFICU) | 49% | Novel Coronavirus<br>Pneumonia Emergency<br>Response<br>Epidemiology<br>Team(38) |
| Proportion of infected individuals requiring intensive care | IFR divided by CFICU |  |
| Length of hospital admission | 10 | Wang et al(39) |
| Length of intensive care unit stay | 7.5 | Assumed |

**Fig. S1.**
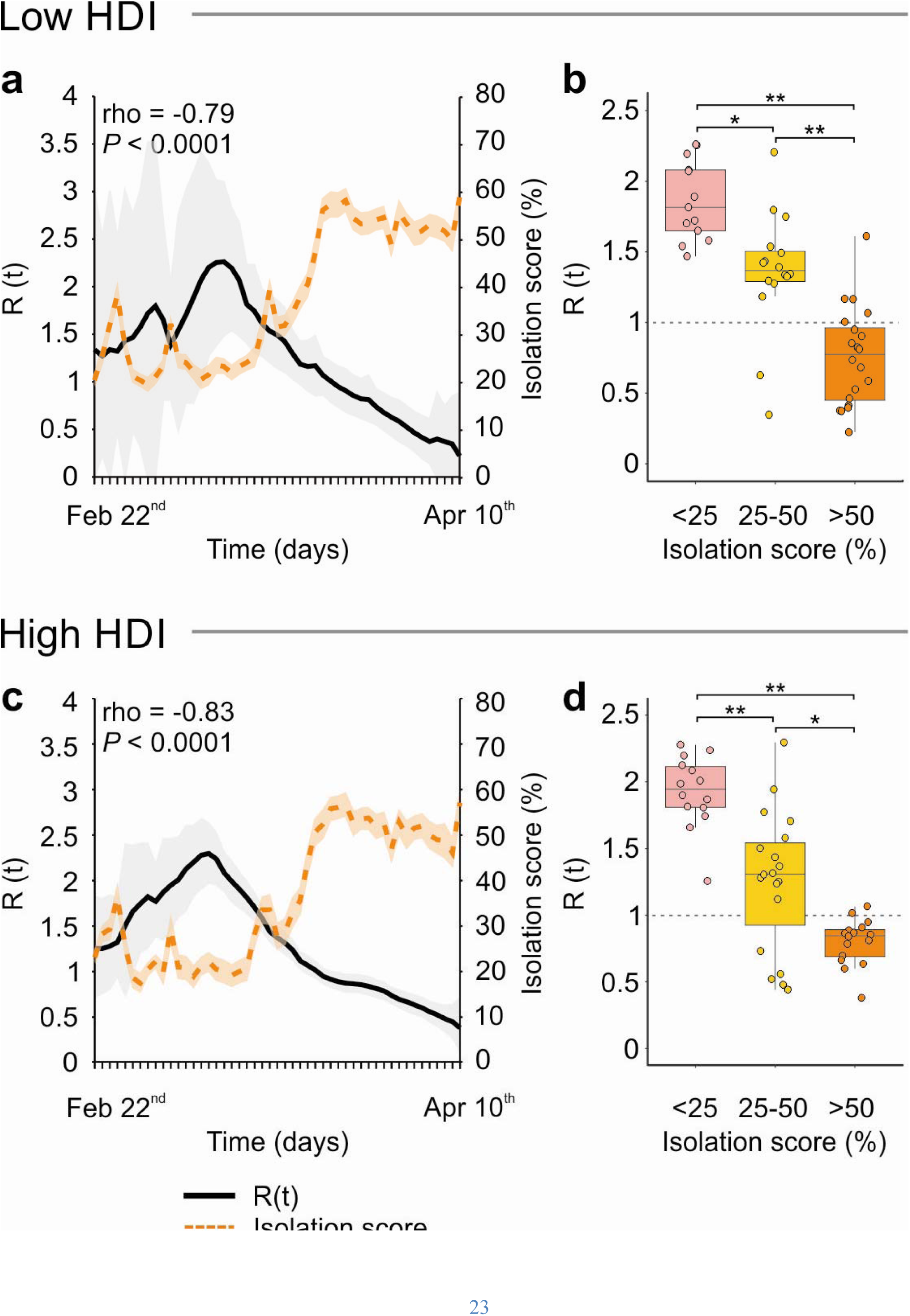
A and C: Time dependent reproductive number (R(t)) and isolation index per day from the cities with lower and higher HDI of Rio and São Paulo, respectively; B and D: boxplot showing the median, interquartile range and range of R(t) stratified by isolation index (<25%, 25–50%, 50%) from cities with lower and higher HDI of Rio and São Paulo, respectively.

